# Safety and Efficacy of Vesemnogene Lantuparvovec, an AAV-Based Gene Therapy in Young Children Under 24 Months with Spinal Muscular Atrophy in Low- and Middle-Income Countries. Second Interim Report (August 2025)

**DOI:** 10.1101/2025.04.13.25325764

**Authors:** Lock Hock Ngu, Qingling Mo, Shiqi Li, Teck Hock Toh, Jia Ni Lee, Kok Chong Lim, Edi Setiawan Tehuteru, Rahmi Lestari, Chinnuwat Sanguansermsri, Hany Abueita, Samson Gwer, Li Li, Zhuowei Wang, Salman Kirmani, Julia X. Chen, Yilia Y. Cai, Nanette N. Zheng, Yanqing Li, Qian Yang, Yanqiong Tang, Yeqing Li, Jason Z. Ye, Silk J. Shi, Jack F. Hong, Amy Y. Chen, Cole K. Zheng, Sanbin Wang, Teck-Onn Lim, Bruce T. Lahn, Austin T. Gao

## Abstract

**Introduction:** Spinal muscular atrophy (SMA) is a monogenic neuromuscular disorder due to the survival motor neuron 1 mutation. Onasemnogene abeparvovec is a U.S. FDA approved single-dose gene therapy for SMA, but it is priced at USD 2.13 million per patient which severely limits its accessibility in low- and middle-income countries (LMICs). We conducted a phase 1 trial on vesemnogene lantuparvovec, an affordably priced substitute to onasemnogene abeparvovec intended for use in LMICs.

**Methods:** Sixteen patients with SMA (eight each with type 1 and type 2) received a single dose of intrathecal vesemnogene lantuparvovec. Eleven patients received a low dose (1.5 × 10^14^ vg) and five received high dose (3.0 × 10^14^ vg). The primary outcomes were safety, and efficacy with the change from baseline in the developmental gross motor milestones achieved according to World Health Organization (WHO) criteria. Overall survival was primarily evaluated in type 1 SMA patients. This trial was registered with ClinicalTrials.gov NCT06288230.

**Results:** As of the data cutoff in August 2025, 16 patients enrolled have been followed-up to at least 3-month post-treatment. The median age at diagnosis and dosing were 4.4 months (0.0, 18.0) and 10.7 months (2.8, 22.5), respectively. The most common adverse event (AE) was transiently increased aspartate aminotransaminase which occurred in 11 patients (69%), with one patient having a level twice the upper limit of normal. No patient required prolonged prednisolone prophylaxis. The most serious AEs were respiratory tract infections which occurred in four (50%) of eight patients with type 1 SMA, leading to invasive ventilation in two and one of them eventually died.

Among the eight patients with type 1 SMA, one had gained one new WHO milestone at 3- month post-treatment. Among the eight patients with type 2 SMA, at 3-month post treatment, three patients had gained at least one new WHO milestone while one had gained three new milestones and could crawl.

**Conclusions:** In patients with type 1 and type 2 SMA below 24 months, a single intrathecal dose of vesemnogene lantuparvovec was safe and well-tolerated, resulting in improved developmental gross motor milestones, which contrast with patients referred to the trial but were untreated. Further studies are necessary to confirm this gene therapy’s long term safety and efficacy.

## Introduction

Spinal muscular atrophy (SMA) is a monogenic neuromuscular disorder due to mutation of the survival motor neuron 1 (*SMN1*) gene resulting in reduced SMN protein. SMA is a relatively common genetic disease (prevalence 1 in 14,694 newborns [1]). Nevertheless it remains a leading cause of infant mortality, especially in low- and middle-income countries (LMICs) despite the advent of disease-modifying drugs such as nusinersen and risdiplam which are costly lifelong treatments.

Onasemnogene abeparvovec is a single-dose gene therapy for SMA. It is particularly suitable for wider application in LMICs where the medical infrastructures to support life-long treatment are limited. It is remarkably effective in presymptomatic infants [2,3] and symptomatic infants less than six months of age [4-7]. Clinical benefits were observed up to age 24 months though its efficacy decreased when given to children with increasing age [8]. The latter is also attested by a recent trial of a single intrathecal dose of onasemnogene abeparvovec in older children up to five years of age [9].

Onasemnogene abeparvovec-xioi (Zolgensma, Novartis) has been approved by the U.S. FDA since 2019 for treating SMA in pediatric patients less than 2 years of age. Unfortunately, it is priced at USD 2.13 million per patient [10], severely limiting its accessibility in LMICs. The availability of a more affordable substitute to onasemnogene abeparvovec will significantly mitigate the health burden posed by SMA in LMICs. Vesemnogene lantuparvovec, an adeno-associated virus (AAV) based gene therapy for SMA, was developed specifically for this purpose. To ensure its affordability in LMICs, vesemnogene lantuparvovec will be priced close to its manufacturing costs, and the technology will be transferred at zero or minimal cost to individual LMICs who wish to adopt this to continue its development leading to local approval.

The vector construct of vesemnogene lantuparvovec is rAAV9.OEE.h-SMN1. Like onasemnogene abeparvovec-xioi, this vector is a nonreplicating, recombinant serotype 9 adeno-associated virus (rAAV9) containing a non-codon-optimized, full-length human survival motor neuron 1 transgene (rAAV9-*SMN1*) under the control of a promoter to restore functional SMN1 protein in motor neurones. The amino acid sequence of the SMN1 protein produced from the transgene is identical to that of normal wild-type hSMN1 protein. In vesemnogene lantuparvovec, the expression cassette flanked by inverted terminal repeats, comprises, in addition to the transgene (human *SMN1* cDNA), an optimized promoter, an engineered post-transcriptional regulatory element, and the bovine growth hormone (BGH) polyadenylation sequence. In onasemnogene abeparvovec-xioi the expression cassette consists of the chicken β-actin promoter, simian virus 40 intron and the BGH polyadenylation sequence too.

Vesemnogene lantuparvovec is currently undergoing clinical trial (NCT06288230) in China and early access use outside China for patients unable to travel to China to be enrolled in the trial. We report here preliminary data on its safety and efficacy in children treated to date (August 2025).

## Methods

This prospective, single arm ascending dose study was carried out at a single center in Kunming China. The study was performed in compliance with the principles outlined in the Declaration of Helsinki. The study site’s institutional review-board approved the study protocol, and all parents or legal guardians gave written informed consent. Because of overwhelming demands for the study treatment, after preliminary safety and efficacy were observed, the study treatment was also available under early access use in LMICs outside China. The study was registered under ClinicalTrials.gov (NCT06288230).

### Study patients

Patients aged 24 months or younger who had genetically confirmed SMA (biallelic deletion of *SMN1*) without the genetic modifier (c.859G>C) [11] were enrolled into the study. Either symptomatic or presymptomatic patients were eligible for enrollment. Key exclusion criteria were contraindications for spinal tap procedure or administration of intrathecal therapy, elevated anti–AAV9 antibody titers >1:50 as determined by ELISA assay, use of invasive ventilatory support (tracheostomy with positive pressure) or non-invasive ventilatory support ≥ 12 hours daily, severe joints contractures and / or scoliosis, active respiratory infection, active viral infection or other severe non–respiratory tract infection, abnormal liver function tests (alanine aminotransferase (ALT) and aspartate aminotransferase (AST) ≥ 2 × upper limit of normal) and other co-morbidities that in the opinion of the investigator may increase the risk of gene replacement therapy. Eligible patients were administered either low dose (1.5 × 10^14^ vg) or high dose (3.0 × 10^14^ vg).

### Study treatment

All patients received the study treatment as follows:

- Oral prophylactic prednisolone (1 mg/kg/day) starting one day prior to intrathecal vesemnogene lantuparvovec dosing and continuing for four weeks. Prednisolone dosing was then tapered off over 2 to 4 weeks if liver function test results were normal.
- A single intrathecal injection of vesemnogene lantuparvovec was given under sedation or anesthesia. Following administration, patients were placed in the Trendelenburg position for 15 minutes to enhance the distribution of the vector to cervical and brain regions. Patients were observed at the hospital for at least 48 hours prior to discharge.

### Study assessments

Safety assessments consisted of monitoring adverse events (AEs) and serious adverse events (SAEs), monitoring hematology and blood chemistry results and physical examinations.

The primary efficacy outcome was the change from baseline in the developmental gross motor milestones achieved according to World Health Organization (WHO) criteria (sitting without support, standing with assistance, hands-and-knees crawling, walking with assistance, standing alone and walking alone) [12]. This outcome measure was captured on video and centrally reviewed. Secondary outcomes were change from baseline in requirements for non-invasive ventilatory support and feeding support via a nasogastric tube or gastrostomy tube.

### Statistical analysis

Safety and efficacy analyses were performed on an intention-to-treat population, including all patients who had received a single dose of vesemnogene lantuparvovec. Safety analysis was evaluated based on the incidence of AEs and SAEs. Efficacy analyses assessed changes in the distribution of WHO motor milestones from baseline to post-treatment, individual trajectories of motor milestone development following treatment, and the proportion of patients achieving at least one WHO motor milestone.

## Results

The first patient was enrolled and treated in May 2024. After the first three patients were treated without overt safety signals, the trial enrollment was expanded to referrals form gobal LMICs. Between October 2024 and August 2025, we received a large number of referrals for enrollment. One hundred twenty-eight patients were screened and assessed for eligibility; 102 were ineligible because they had previously been referred for palliative care (n = 16), were tested AAV antibody positive (n = 4), or were aged above 24 months. There remained 26 eligible patients. Treatment capacity was constrained by the limited number of doses produced in the clinical trial batch. By August 2025, only 16 patients had received treatment, whereas five had died awaiting treatment and another five patients remained on the waiting list. All 16 treated patients had completed follow-up to 3-month post treatment, and none had withdrawn from the study.

Of the 16 treated patients, five were from Vietnam, four from Pakistan, two from Palestine, and one each from Malaysia, Thailand, Indonesia, India and Kenya. **Table 1** shows the baseline characteristics of the patients. The median age at treatment administration was 10.7 months (range 2.8-22.5). Eight patients have two *SMN2* copies, eight have three copies. Eight patients were identified as having type 1 SMA, characterized by diagnosis before 12 months of age, symptom onset prior to 6 months, and two copies of the *SMN2* gene. The other eight patients had type 2 SMA. Eleven patients had received risdiplam treatment; none had nusinersen. At baseline, all patients could not sit unsupported except for three sitters and two were able to stand with assistance. All patients with type 1 SMA were unable to sit unsupported except for one sitter. Among patients with type 1 SMA, one had required feeding via nasogastric tube and another required feeding via gastronomy tube. One patient was on invasive ventilation though three had required intermittent non-invasive ventilatory support using a bilevel positive airway pressure machine (BiPAP) during previous episodes of respiratory infection. No patients with type 2 SMA had required tube feeding or BiPAP.

**Table 1.**
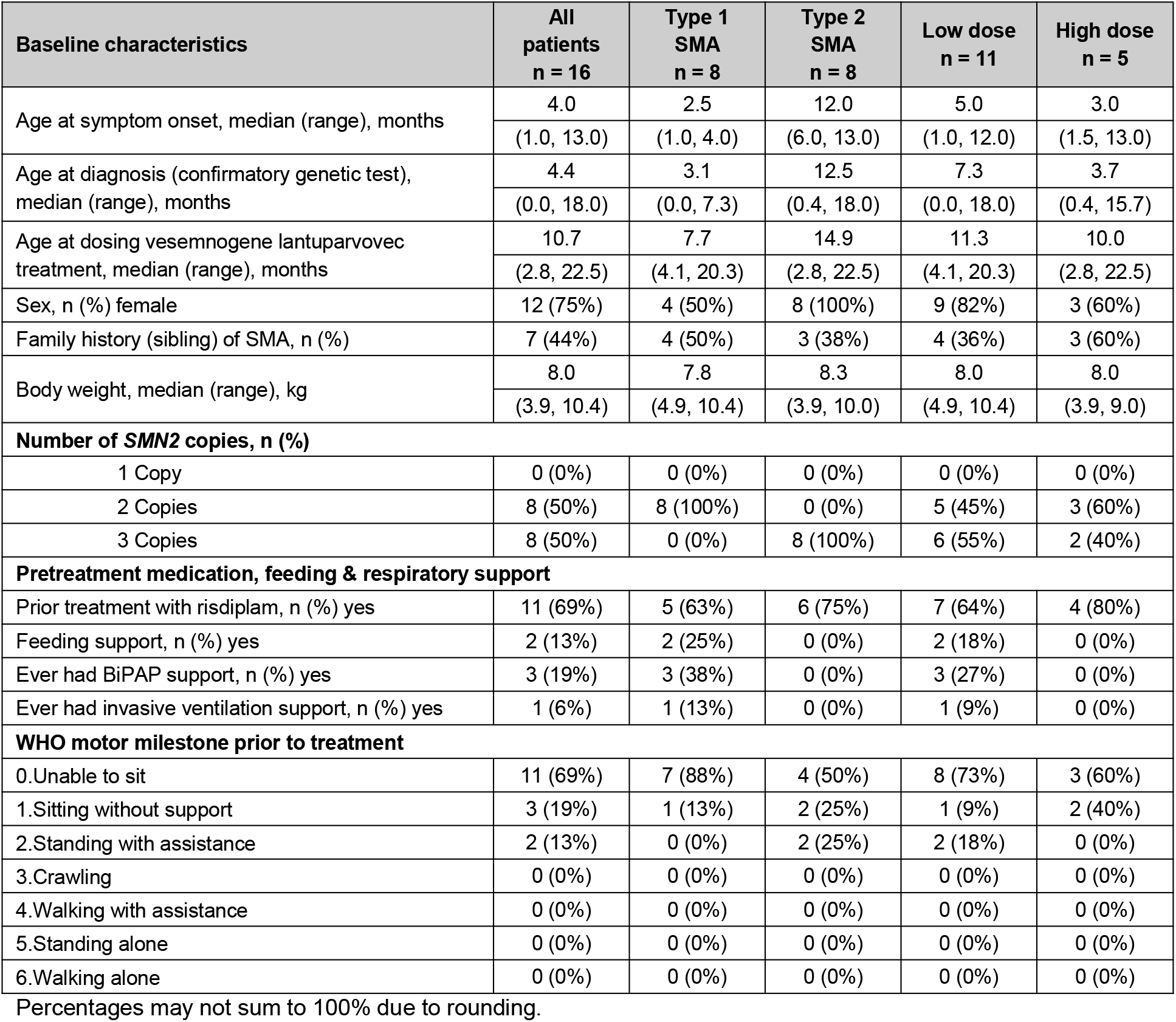
Baseline characteristics of patients.

### Safety results

Ninety-six adverse events (AEs) were reported in the trial to date, with 53 events occurring among patients with type 1 SMA and 43 in type 2 SMA.

**Table 2** summarizes all the reported AEs. The most common AE was increased AST (11 events, 69%), with one patient showing levels more than 2 × upper limit of normal. Other events werelymphocyte count increased, low density lipoprotein increased, pyrexia (5 events each, 31%), lower respiratory tract infection, decreased fibrinogen, increased lactate dehydrogenase (4 events each, 25%).

**Table 2.** Incidence of AEs.

| AE Terms | All patients<br>n = 16 | Low dose<br>$1.5 \times 10^{14}$ vg<br>n = 11 | High dose<br>$3.0 \times 10^{14}$ vg<br>n = 5 |
| --- | --- | --- | --- |
|  | n (%) | n (%) | n (%) |
| Any AEs | 15* (94%) | 11 (100%) | 4 (80%) |
| Pyrexia | 4 (25%) | 2 (18%) | 2 (40%) |
| Rash | 2 (13%) | 2 (18%) | 0 (0%) |
| Blood Mycoplasma pneumonia antibody positive | 1 (6%) | 1 (9%) | 0 (0%) |
| Lower respiratory tract infection | 4 (25%) | 4 (36%) | 0 (0%)** |
| Respiratory difficulty requiring ventilation | 4 (25%) | 4 (36%) | 0 (0%)** |
| Respiratory failure requiring invasive ventilation | 2 (13%) | 2 (18%) | 0 (0%)** |
| Feeding support | 5 (31%) | 5 (45%) | 0 (0%) |
| Aspartate aminotransferase increased >ULN | 11 (69%) | 7 (64%) | 4 (80%) |
| Aspartate aminotransferase increased >2x ULN | 1 (6%) | 1 (9%) | 0 (0%) |
| Alanine aminotransferase increased >ULN | 3 (19%) | 2 (18%) | 1 (20%) |
| Alanine aminotransferase increased >2x ULN | 0 (0%) | 0 (0%) | 0 (0%) |
| Troponin I increased | 1 (6%) | 1 (9%) | 0 (0%) |
| Thrombocytopenia | 0 (0%) | 0 (0%) | 0 (0%) |
| Sensory abnormalities | 0 (0%) | 0 (0%) | 0 (0%) |

Percentages may not sum to 100% due to rounding.
Abbreviation: AEs, adverse events; ULN, upper limit of normal.
\*Ninety-six AEs occurred among 15 patients; some patients have more than one event while 1 had none at all.
\*\*Safety data for patients on high dose observed to 1-month post treatment to date.

Elevated AST, ALT, and troponin I were reported as related to the study treatment. All elevated AST events resolved spontaneously. All patients had received prednisolone prophylaxis; no patient required prolonged prednisolone prophylaxis after the prescribed four weeks duration.

Most SAEs were respiratory tract infections that occurred in four (50%) of eight patients with type 1 SMA and one (13%) of eight patients with type 2 SMA. These four patients with type 1 SMA had led to hypoxemia requiring ventilatory support, two of whom had progressed to require invasive ventilation and one of whom died at 6-month post-treatment. These events were considered related to the natural history of type 1 SMA. No patients with type 2 SMA required any ventilatory support.

No signs or symptoms of thrombotic microangiopathy or sensory abnormalities were reported, although electrophysiological testing was not conducted routinely in these young children.

### Efficacy results

All parents reported improvement in the motor function of their babies within days to weeks after vesemnogene lantuparvovec treatment. Reported improvement in motor movement not previously observed in these babies were eyes and head able to follow moving objects, hand grasping object, holding head erect without support, rolling from back to sides, and vocalizing “papa, mama”.

**Table 3** shows the distribution of WHO motor milestones at baseline and post-treatment and **Figure 2** depicts the longitudinal trajectory in individual patients. At 3-month post-treatment, 4 patients (25%) had gained at least one new WHO milestone, one with three new WHO milestones and were able to crawl.

**Table 3.** WHO motor milestones at baseline and achieved post-treatment.

| WHO motor milestone | Baseline | 1-month post-treatment | 3-month post-treatment | 6-month post-treatment |
| --- | --- | --- | --- | --- |
|  | n = 16 | n = 16 | n = 16 | n = 9* |
| Unable to sit | 11 (69%) | 8 (50%) | 9 (56%) | 4 (44%) |
| Sitting without support | 3 (19%) | 6 (38%) | 4 (25%) | 1 (11%) |
| Standing with assistance | 2 (13%) | 2 (13%) | 0 (0%) | 1 (11%) |
| Crawling | 0 (0%) | 0 (0%) | 3 (19%) | - |
| Walking with assistance | 0 (0%) | 0 (0%) | 0 (0%) | 2 (22%) |
| Standing alone | 0 (0%) | 0 (0%) | 0 (0%) | - |
| Walking alone | 0 (0%) | 0 (0%) | 0 (0%) | - |

Percentages may not sum to 100% due to rounding.
\*: One patient died at 6-month post-treatment.

**Figure 1.**
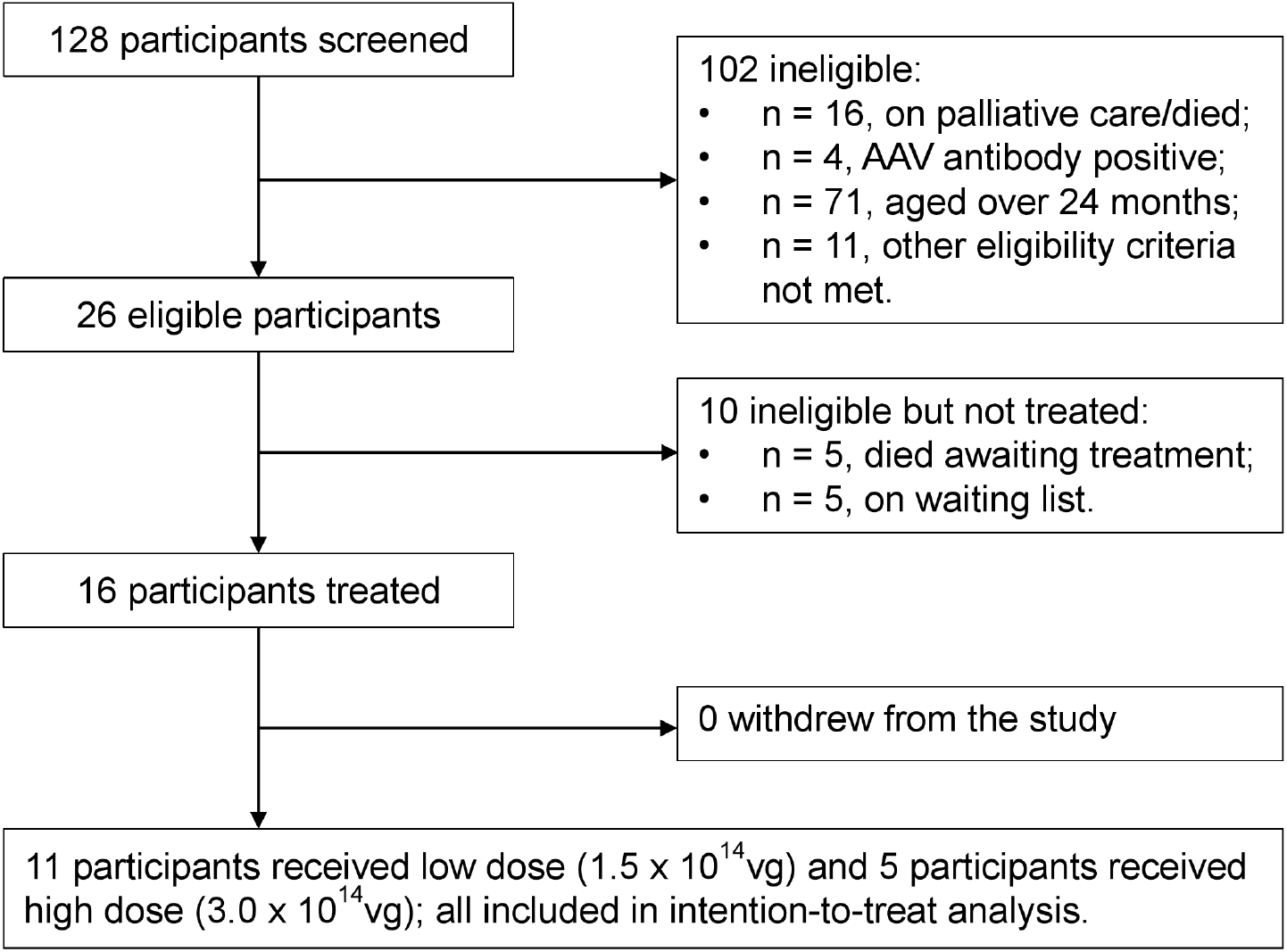
Trial Profile.

**Figure 2.**
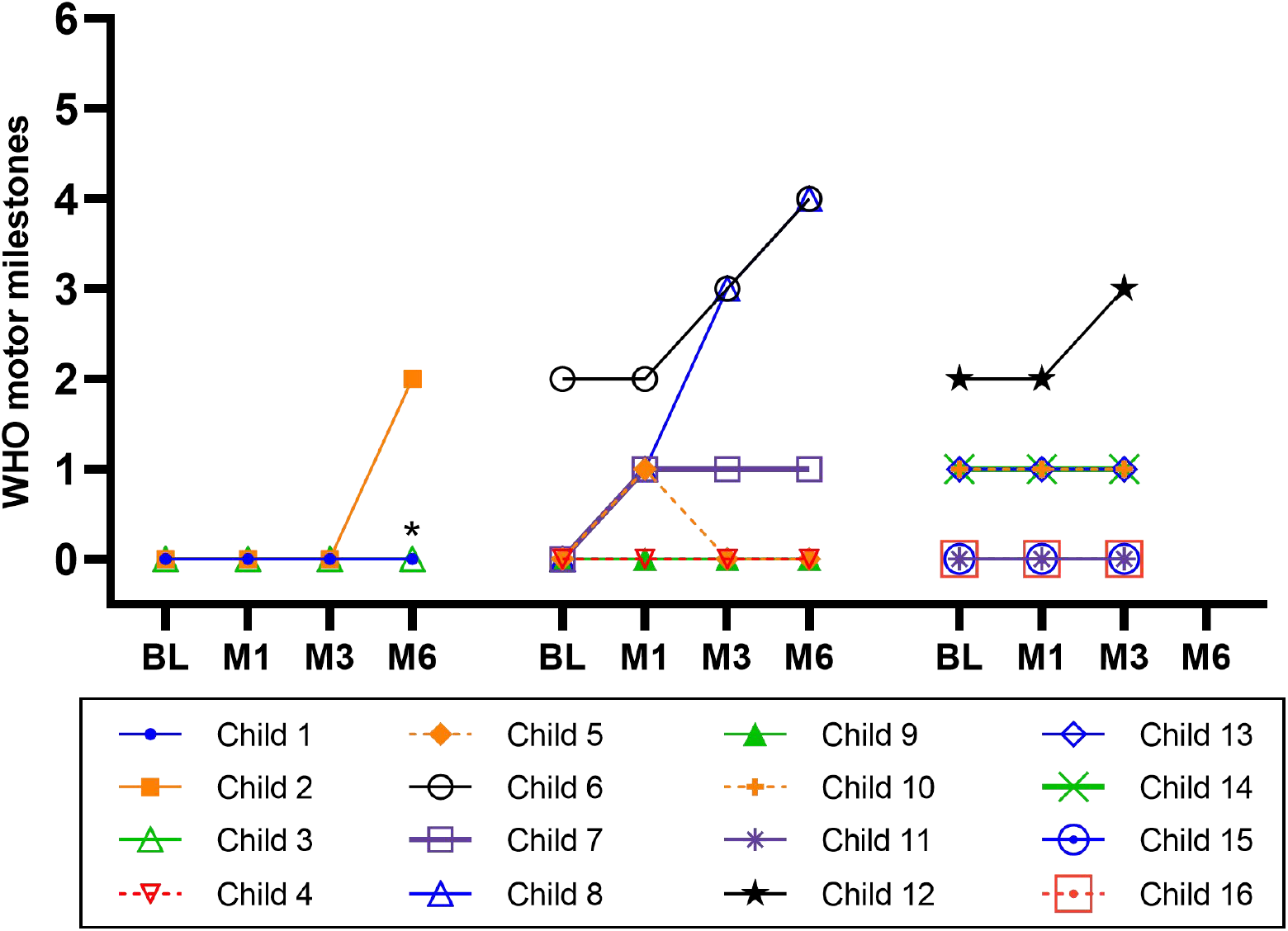
Trajectory of WHO motor milestones achieved post-treatment among the study patients. Abbreviations: BL, baseline; M1, 1 month; M3, 3 months; M6, 6 months post-treatment. WHO motor milestones: 0, Unable to sit; 1, Sitting without support; 2, Standing with assistance; 3, Crawling; 4, Walking with assistance; 5, Standing alone; 6, Walking alone. *, Child 1 died in M6.

Among the eight patients with type 1 SMA, only 1 had gained one new WHO milestone at 3-month post-treatment (**Table 4**). As of the data cutoff in August 2025, three (38%) had required nasogastric tube placement and four patients (50%) had required non-invasive ventilatory support in the course of their respiratory infections, two (25%) of them had progressed to require invasive ventilation, one of them died at 6-month post-treatment.

**Table 4.** WHO motor milestones at baseline and achieved post-treatment by type of SMA.

| WHO motor milestone | Type 1 SMA |  |  |  | Type 2 SMA |  |  |  |
| --- | --- | --- | --- | --- | --- | --- | --- | --- |
|  | Baseline | M1 | M3 | M6 | Baseline | M1 | M3 | M6 |
|  | n = 8 | n = 8 | n = 8 | n = 5* | n = 8 | n = 8 | n = 8 | n = 4 |
| Unable to sit | 7 (88%) | 6 (75%) | 6 (75%) | 3 (60%) | 4 (50%) | 2 (25%) | 3 (38%) | 1 (25%) |
| Sitting without support | 1 (13%) | 2 (25%) | 2 (25%) | 1 (20%) | 2 (25%) | 4 (50%) | 2 (25%) | - |
| Standing with assistance | 0 (0%) | 0 (0%) | 0 (0%) | - | 2 (25%) | 2 (25%) | 0 (0%) | 1 (25%) |
| Crawling | 0 (0%) | 0 (0%) | 0 (0%) | - | 0 (0%) | 0 (0%) | 3 (38%) | - |
| Walking with assistance | 0 (0%) | 0 (0%) | 0 (0%) | - | 0 (0%) | 0 (0%) | 0 (0%) | 2 (50%) |
| Standing alone | 0 (0%) | 0 (0%) | 0 (0%) | - | 0 (0%) | 0 (0%) | 0 (0%) | - |
| Walking alone | 0 (0%) | 0 (0%) | 0 (0%) | - | 0 (0%) | 0 (0%) | 0 (0%) | - |

Abbreviations: M1: 1 months post-treatment, M3: 3 months post-treatment, M6: 6 months post-treatment, M9: 9 months post-treatment, M12: 12 months post-treatment.
\*: One patient died at 6-month post-treatment.

Among the eight patients with type 2 SMA at 3-month post treatment, three patients had gained at least one new WHO milestone while one had gained three new milestones and could crawl (**Table 4**). As of the data cutoff in August 2025, no patients with type 2 SMA had required any ventilatory or feeding support.

**Table 5** compares the distribution of WHO motor milestones between patients treated using low dose (1.5 × 10^14^ vg) and high dose (3.0 × 10^14^ vg). At 3-month post treatment, there is no apparent difference in the distribution of WHO motor milestones achieved post-treatment between these two groups.

**Table 5.**
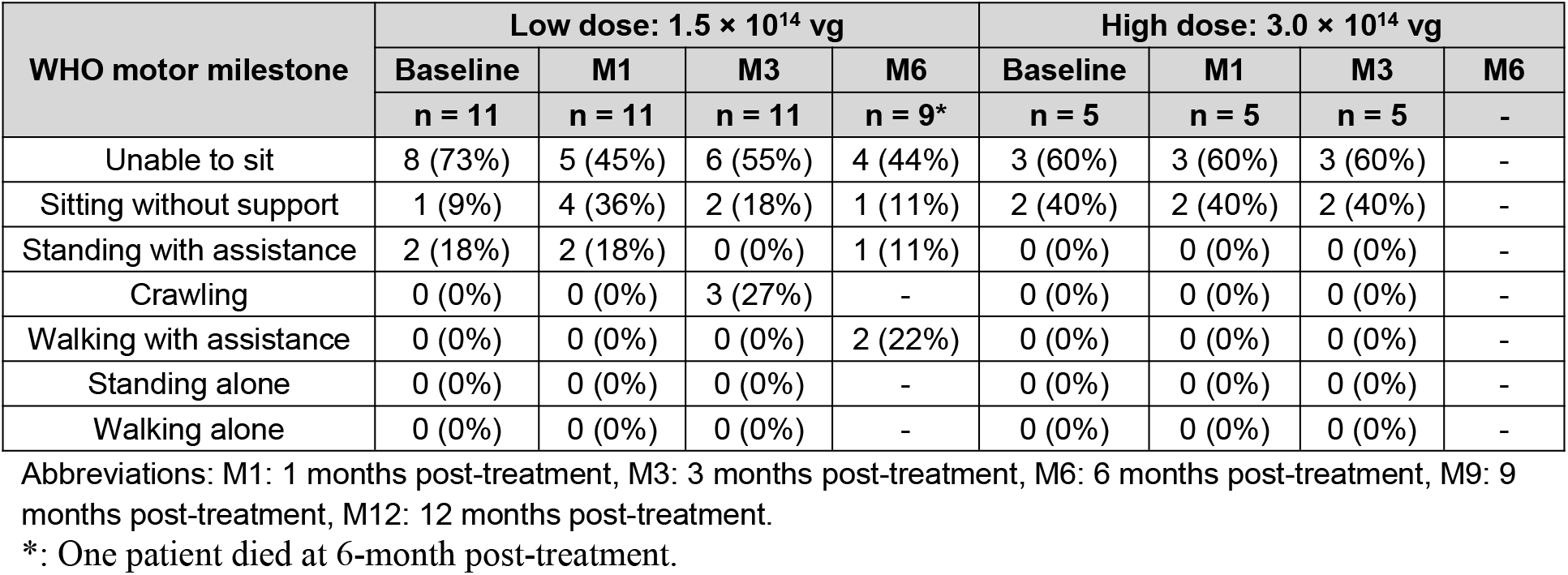
WHO motor milestones at baseline and achieved post-treatment by dosing level.

## Discussion

We conducted a phase 1 study of vesemnogene lantuparvovec for treating children with SMA enrolled from diverse LMIC settings across populations and geographies.

Acute AEs are expected to occur early from previous experience of AAV based gene therapy trials. The most concerning are immune-mediated toxicities associated with AAV, targeting in particular the liver and heart. Consistent with safety data from the use of AAV-based gene therapy for SMA, elevated transaminases occurred in the majority (75%) of patients within one month after receiving vesemnogene lantuparvovec. In contrast to the experience with onasemnogene abeparvovec administered intravenously, elevated transaminases observed in this trial were isolated events without associated signs / symptoms and the elevated levels resolved spontaneously in all patients. Only one patient had elevated AST at twice the normal upper limit which resolved within a week. No patient required prolonged prednisolone prophylaxis. Increased blood troponin I occurred in only 1 patient. Like elevated transaminase, this was also an isolated event without associated signs and symptoms. Overall, intrathecal administration of a single dose of vesemnogene lantuparvovec was safe and well-tolerated, consistent with the safety profile observed in the first trial of onasemnogene abeparvovec administered intrathecally [9].

On the efficacy of vesemnogene lantuparvovec, we have limited data on only 16 patients observed up to 3 to 6 months post treatment to date. Four patients (25%) had gained at least one new WHO milestone at 3-month post treatment. Remarkably, one patients had gained three new WHO milestones in three months and were able to crawl.

Nevertheless, this is a single-arm phase 1 study. In the absence of a comparable control group, because of the possibility of confounding bias, it is not possible to infer whether the improved motor development is due to developmental maturation or to the effect of vesemnogene lantuparvovec.

- Randomized placebo control trial is ideal but have been deemed ethically problematic even in LMIC settings where palliation is the standard of care for infants and toddlers with type 1 SMA.
- It is also practically impossible to assemble a natural history cohort in this setting. Patients were enrolled from diverse LMIC settings with large variations in the local standard of care. In many LMICs, genetic tests for SMA are not available, infants with flaccid weakness and progressive respiratory failure are routinely referred to palliative care without genetic diagnosis of underlying disease, and there is variable access to charity-funded risdiplam among countries. Hence, patients referred for this trial are a highly selected population and constitute a fraction of all incident SMA by virtue of actually being diagnosed SMA, and 69% were treated with risdiplam.
- Patients most comparable to the treated patients in this trial are those who were enrolled but for various reasons were not treated. There are 2 such groups: (1) Patients with Type 1 SMA who were tested AAV antibody positive and therefore ineligible for trial. Of three such patients, two have progressed to respiratory failure requiring invasive ventilation within three months after referral for trial; (2) Patients with type 1 SMA who were on waiting list. Delay in getting to the study treatment was inevitable because parents and their doctors took time to consider the trial treatment option, delay in genetic and AAV antibody tests which require shipment of blood samples to the central lab, delay in obtaining passport / visa to travel to China to enter the trial. Of seven such patients, five had died within six months after referral for trial. The rapidly progressive adverse outcomes among these referred but untreated patents contrast with those treated in the trial. Further among the eight patients with type 2 SMA given study treatment but were not at risk of respiratory failure or death; three patients had gained at least one new WHO milestone while one had gained three new milestones and could crawl. This finding is consistent with previous studies on AAV-based gene therapy which have shown rapid onset of action [9], even among patients who had prior disease modifying treatment [13]. Similar rapid and marked improvement have not been reported among patients on waiting list including those on risdiplam.

This study has several limitations

- As mentioned above, the absence of a randomized control group and the use of external control raise the possibility of confounding bias. This is a problem common to all intervention studies in rare diseases when randomization is not practically or ethically feasible [14].
- The sample size is small for a phase 1 study. Later phase studies will be initiated in LMICs to support local registration. The technology transfer of vesemnogene lantuparvovec to these LMICs is currently ongoing.
- The duration of observation to date is limited to 3 to 6 months after vesemnogene lantuparvovec treatment. Longer-term safety and efficacy assessment will require observations on a larger sample of patients for longer duration.

In conclusion, this phase 1 trial has provided preliminary evidence that the intrathecal administration of vesemnogene lantuparvovec, a novel AAV9-based gene therapy, is safe and also well-tolerated in young children with type 1 and type 2 SMA in LMIC settings. Despite the small cohort and limited follow-up duration, early post-treatment gains in WHO motor milestones, particularly in patients with type 2 SMA, are promising. Importantly, these improvements were achieved in a population with limited access to standard-of-care therapies and in a setting where gene therapy has historically been inaccessible due to cost barriers. The absence of a randomized control group and the study population’s heterogeneity limit the efficacy conclusions’ strength. However, the stark contrast in the rapid progression and high mortality observed among untreated referred patients underscores the potential benefits of vesemnogene lantuparvovec. These data reinforce the requirement for cost-efficient, scalable gene therapy solutions within LMICs as a priority matter. Further studies with larger sample size, extended follow-up, and locally contextualized registries are warranted to validate these findings and support regulatory approval and widespread adoption. The ongoing technology transfer and local manufacturing efforts are important steps towards sustainable SMA care and universal global access to innovative gene therapy.

## Data Availability

All data produced in the present study are available upon reasonable request to the authors

